# Attention-Enhanced U-Net Segmentation for Reliable Detection of Circulating Tumor-Associated Cells

**DOI:** 10.64898/2026.03.07.26347846

**Authors:** Massimo Cristofanilli, Sewanti Limaye, Nitesh Rohatgi, Timothy Crook, Humaid Al-Shamsi, Andrew Gaya, Raymond Page, Aditya Shreeniwas, Darshana Patil, Vineet Datta, Dadasaheb Akolkar, Stefan Schuster, Prashant Agrawal, Shoeb Patel, Pradyumna Shejwalkar, Snehal Golar, Ajay Srinivasan, Rajan Datar

**Author notes:** Corresponding Author: Massimo Cristofanilli.

## Abstract

**Background:** Circulating tumor associated cell (CTAC) detection-based multi-cancer early detection (MCED) strategies may be hindered by the rarity of CTACs among millions of peripheral blood nucleated cells (PBNCs). We developed an advanced U-Net-based encoder-decoder model for pixel-level CTAC discrimination that integrates attention-gated skip connections to preserve morphological and fluorescence details.

**Methods:** Model suitability was explored in an initial cohort of asymptomatic individuals (n = 428) and patients with advanced solid tumors (n = 354). A case-control study assessed clinical performance in therapy-naive stage I/II cancer patients (n = 185), individuals with benign conditions (n = 129), and asymptomatic individuals (n = 111). The model was then validated across four prospective studies on distinct populations: recurrent cancer cases with low tumor burden (n = 224); patients with solid tumors in the peri-operative setting (n = 17); suspected cancer cases (n = 259); and asymptomatic individuals (n = 7,183), respectively. All studies used blinded peripheral blood specimens from which PBNCs were isolated, stained for EpCAM / Hoechst 33342, and imaged. Ground truth annotations were established via pathologist review. The U-Net pipeline encoded spatial information in the images via convolutional and pooling layers and generated pixel-wise segmentation masks to identify CTACs. In all studies, sensitivity was based on CTAC detection rate in cancer specimens and CTAC undetectability rate in specimens from healthy asymptomatic individuals or those with benign conditions

**Results:** In the exploratory study, the model had 90.68% (95% CI: 87.16%, 93.50%) sensitivity and 99.53% (95% CI: 98.32%, 99.94%) specificity. In the case-control cohort, the model had 88.65% sensitivity (95% CI: 83.17%, 92.83%), 78.95% (95% CI: 71.03%, 85.53%) specificity in benign conditions, and >99.9% specificity in asymptomatic individuals. Among the four prospective studies, the model had: (a) 91.96% (95% CI: 87.60%, 95.17%) sensitivity in pretreated patients with low tumor burden; (b) 100% sensitivity in pre-surgery specimens, and 29.41% sensitivity in post-surgery specimens; (c) 96.34% PPV (95% CI: 93.22%, 98.05%) and a 32.35% NPV (95% CI: 25.58%, 39.95%) for diagnostic triaging; and, (d)11% PPV (95% CI: 31.72%, 53.24%) and 99.97% NPV (95% CI: 99.90%, 99.99%) for MCED in healthy asymptomatic individuals.

**Conclusions:** The attention-enhanced U-Net achieved robust, generalizable performance for CTAC-detection in case-control and prospective cohorts, supporting its clinical utility for accurate cancer detection.

## Introduction

Circulating tumor cells (CTCs) are cancer cells that detach from malignant tumors, enter the bloodstream, and potentially lead to distant metastasis [1,2]. CTCs have been recognized as potential biomarkers for early cancer detection [3], disease monitoring[4], and treatment response assessment[5]. CTCs are clinically relevant analytes and their detection and characterization may aid cancer diagnostics, prognostication, and treatment monitoring[6,7]. We have previously described circulating tumor associated cells (CTACs) that include classical CTCs, and other tumor-associated cells that are positive for epithelial markers, irrespective of CD45 status[8]. Assessing CTACs rather than only CTCs improves sensitivity by capturing epithelial-marker–positive cells that may be missed by CD45-restricted definitions, thus better reflecting tumor heterogeneity and tumor–immune interactions that drive tumorigenic mechanisms. Consequently, CTACs provide a comprehensive of circulating tumor biology. We have also described the process for identification of CTACs via immunostaining followed by image analysis[9]. Conventional CTAC detection via immunostaining along with microscopy or flow cytometry face sensitivity limitations in addition to labor-intensive protocols, and varied specificity due to subjective manual interpretation[10,11].

Artificial intelligence (AI) based automation of cell detection and classification can qualitatively and quantitatively improve medical image analysis [12,13]. However, conventional convolutional neural networks (CNNs) often struggle with the fine-grained discrimination required for identifying rare CTACs among the exponentially higher background of normal (i.e., non-malignant) peripheral blood nucleated cells (PBNCs) [14]. This challenge is compounded by subtle differences in morphological and marker expression characteristics among CTACs, that can confound accurate differentiation of CTACs from nonmalignant cells [15].

The U-Net architecture, originally developed for biomedical image segmentation[16], offers a solution to the skewed signal : noise ratio in imaging. Its encoder-decoder structure with skip connections preserves spatial information while enabling deep feature extraction, making it particularly suitable for tasks requiring pixel-level precision[17]. Despite these advantages, the application of U-Net architectures to CTAC detection from fluorescence microscopy images remains underexplored.

We developed and validated a specialized U-Net-based deep learning pipeline for automated CTAC detection from PBNCs stained with epithelial cell adhesion molecule (EPCAM) and Hoechst 33342. Our pipeline emphasizes pixel-level segmentation, preserving morphological and fluorescence details that are essential for accurate CTAC identification. By integrating contextual information and employing skip connections, our approach overcomes the limitations of traditional CNNs, which often lose fine-grained details through successive pooling operations.

In this manuscript, we present a comprehensive evaluation of our U-Net-based CTAC detection pipeline. We explore the architectural advantages of our customized U-Net design and discuss its potential clinical applications. We assessed the performance characteristics of the process in controlled clinical study cohorts comprising healthy volunteers, individuals with non-malignant conditions, and patients with early-stage cancers. We show robust performance of the process in a prospective patient-screening population.

## Materials and Methods

### Study Design and Subjects

All studies and sub-studies were conducted after approval by the Institutional Ethics Committee (IEC) of the Study Sponsor (Datar Cancer Genetics, DCG), and in accordance with applicable regulatory requirements and the Declaration of Helsinki. Study procedures included the collection of biological specimens (peripheral blood) from (a) asymptomatic individuals with no prior history of cancer diagnosis, (b) individuals with histopathologically confirmed non-malignant (benign) conditions, (c) individuals with histopathologically confirmed cancers, and, (d) individuals with symptoms, or clinical laboratory or radiological findings suspected of cancer. No individual received any treatment or underwent any invasive procedures solely for the purpose of this study – any treatments or invasive procedures referenced in this study were a part of the respective standard of care (SoC) management pathways. Informed consent was obtained from all study participants for the collection and use of deidentified specimens for research purposes, and the publication of deidentified data and findings. In addition, the study also evaluated leftover blood specimens from individuals who had previously utilized study sponsor’s oncology diagnostic services (e.g., Exacta, TruCheck / EasyCheck, TruBlood, CancerTrack, ChemoScale) and had consented to the use of leftover deidentified specimens for research and scientific purpose, and the publication of deidentified data and findings; the study sponsor’s test order forms (TOF) include an IEC-approved (optional) consent statement for the research use of deidentified leftover specimens and publication of deidentified data.

### Benign and Confounding Condition Representation

To minimize spectrum bias and evaluate potential sources of false positive results, the study design incorporated a cohort of individuals with benign clinical conditions and non-malignant diagnoses. These included subjects with benign tumors and non-neoplastic conditions that are commonly encountered during real-world cancer diagnostic workups. Such conditions can produce circulating cellular or nucleic acid signals that may mimic malignancy and therefore represent critical confounders in the evaluation of multi-cancer early detection (MCED) technologies. The benign cohort comprised individuals diagnosed with conditions including benign tumors, inflammatory diseases, autoimmune disorders, and other non-malignant pathological states. Where available, clinical diagnoses were confirmed through standard-of-care investigations including imaging, laboratory testing, and histopathological evaluation. To further reflect real-world diagnostic pathways, the suspected-cancer cohort included individuals undergoing clinical evaluation for possible malignancy prior to histopathological confirmation. Blood samples were collected prior to definitive diagnostic biopsy or surgical intervention wherever feasible. This design enabled evaluation of assay performance in a pre-diagnostic clinical setting (“intent-to-test population”), which more closely reflects the intended clinical application of MCED technologies.

### Mitigation of Spectrum and Verification Bias

Recognizing the methodological limitations frequently encountered in case–control validation studies of MCED assays, the present study incorporated both retrospective case–control cohorts and prospective cohorts reflecting real-world clinical use scenarios. Specifically, (a) a case–control cohort including cancer patients and individuals with benign conditions was used to establish analytical performance characteristics, (b) a suspected-cancer cohort representing an intent-to-test population undergoing diagnostic evaluation was included to assess performance in a pre-diagnostic clinical context, and, (c) a large asymptomatic screening cohort was included to explore potential application in population-level cancer detection. Although not all screening participants underwent confirmatory diagnostic procedures, individuals with positive assay findings were advised to undergo further clinical evaluation and follow-up. This approach reflects pragmatic real-world implementation and acknowledges the potential for verification bias inherent in screening studies.

### Sample Collection and Processing

Peripheral blood samples (∼6 mL) were collected in EDTA vacutainers from all participants by venous draw. All samples were processed within 72 hours of collection. Initially, red blood cells (RBC) were lysed using a commercial lysis buffer (BD Pharm Lyse™, BD Biosciences) according to the manufacturer’s instructions. Peripheral blood nucleated cells (PBNCs) were isolated by centrifugation at 400×*g*, 5 min, room temperature (RT). The cell pellet was washed twice with 1x phosphate-buffered saline (PBS) and resuspended in 1 ml of 1x PBS. Cells were fixed with 4% paraformaldehyde (15 min, RT). Immuno-staining of epithelial cell adhesion molecule (EpCAM) was performed using Alexa Fluor 565-conjugated recombinant human anti-CD326 (Clone REA764_Miltenyi Biotech,). Immunostained cells were mounted on adhesive slides (Superfrost Plus, Thermo Scientific) and cover-slipped using anti-fade mounting medium (ProLong Gold, Invitrogen). High-resolution fluorescence microscopy images were acquired using an automated Zeiss Axio Imager 2 slide scanner with oil immersion objective, using Hoechst 33342 and Alexa Fluor-565 (EPCAM) channels at specified wavelengths, and Z-stack acquisition. For each sample, multiple non-overlapping fields were captured, processed as maximum intensity projections, and stored as uncompressed TIFF files to preserve the dynamic range. All samples were processed in batches with positive and negative controls. All laboratory personnel were blinded to the clinical status of the samples during processing.

### Reference Controls

Contrived samples were generated using MCF-7 (breast carcinoma), SW-620 (colorectal adenocarcinoma), SKBR3 (breast carcinoma), A-549 (lung carcinoma), PL-45 (pancreatic cancer), OVCAR-3 (ovarian cancer), and, PC-3 (prostate cancer) cell lines, all of which have well-characterized epithelial features and EpCAM expression (verified periodically by immunostaining). All cell lines undergo periodic mycoplasma testing and short tandem repeat (STR) profiling. Cells were cultured and harvested using standard techniques. Contrived specimens were generated by spiking 100–200 cells into 5 mL of healthy donor whole blood and immunostained (and imaged) as described in the previous section. High-resolution images of biomarker-positive cells were used in initial model development and training, and in analytical validation. Contrived specimens were processed in parallel with study samples according to the established protocols and served as positive controls. The details of control cell lines and critical process reagents are provided in Supplementary Table 1.

### Ground Truth Annotation

To establish ground truth for model training and evaluation, a subset of images was manually annotated by a panel of pathologists. The first step of annotation involved an independent review of images by each pathologist using an in-house annotation software. In the next step, cellular boundaries were manually marked at the pixel-level. Finally, the cells were classified as either CTACs or non-CTACs based on: (a) EPCAM positivity (fluorescence signal localized to cell membrane), (b) Nuclear morphology (irregular nucleus on Hoechst 33342 staining), (c) Cell size (diameter >10 μm), and (d) nuclear-to-cytoplasmic ratio (N:C ≥ 0.7). Annotations with at least 90% agreement were considered in the final ground truth dataset. For discordant observations the consensus was reached through joint review. A total of 2200 cells were annotated, including 1000 confirmed CTACs from cancer patients and 1200 non-CTAC cells from both cancer patients and asymptomatic individuals.

### U-Net Architecture and Implementation

The custom U-Net was designed for detecting CTACs in dual-channel fluorescence images generated from EPCAM / Hoechst 33342 stained specimens. The network processes 512×512-pixel inputs through an encoder-decoder framework with skip connections. The encoder uses 4 blocks of dual 3×3 convolutions (with rectified linear units, ReLU, and batch normalization) followed by 2×2 max pooling, that expands the total channels from 64 to 1024. A bottleneck with two convolutions and dropout (0.5) provides regularization. The decoder mirrors this structure with 4 blocks of transposed convolutions, skip connections, and dual convolutions, reducing channels from 512 to 64. A final 1×1 convolution with sigmoid activation generates pixel-wise CTAC probability maps. In this model, the attention gates in the skip connections highlight relevant spatial features before decoder concatenation[18,19]. Here, the attention coefficient (*α^l^*) was computed as 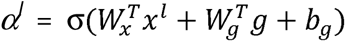, where, ‘*x^l^’* is the feature from the encoder at layer ‘*l’*, *g* is the gating signal from the decoder, 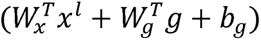 is the linear combination for attention, and ‘σ’ is typically a sigmoid function that yields attention coefficients between 0 and 1. Next, the encoder features were reweighted via element-wise multiplication, as 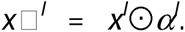. In this manner, the attention gate emphasizes image regions that are more likely to contain CTAC signals, while simultaneously down-weighting regions with noise (e.g., intact PBNCs or cellular debris); *i.e.*, the model focuses its segmentation effort on higher CTAC-probability regions.

### Model Training

Model Training was performed using large image datasets generated from contrived specimens, partitioned into training (70%), validation (15%), and testing (15%) subsets, maintaining stratified representation of CTAC-like cells. The extreme class imbalance (i.e., 1 CTAC per ≥10^6^ blood cells) was addressed by applying a weighted loss approach, coupled with extensive data augmentation. Model training employed a hybrid objective, combining binary cross-entropy and Dice loss (α=0.3), optimized using Adam (initial learning rate = 10^-4^; decay factor 0.9 every 5 epochs). Each batch comprised 16 image patches, with early stopping (patience=15) guided by validation loss. Hyperparameters, including learning rate, weight decay, dropout rate, batch size, and loss-weighting were optimized through Bayesian optimization with 5-fold cross-validation. To improve convergence, we applied cosine annealing with warm restarts [20], starting with 10-epoch cycles and doubling thereafter. Our augmentation pipeline combined both class-aware and generic transformations. Targeted augmentations included CTAC-weighted mix-up (α=0.2), Focal loss (γ=2.0) for hard examples, generation of synthetic CTACs from conditional GANs (tripling minority class samples), random background erasing, and CTAC-instance switching across backgrounds [21,22,23]. GAN-generated CTACs were randomly assigned for validation by pathologists to ensure their synthetic features were biologically plausible and did not negatively impact model generalizability. Generalization was further enhanced through standard augmentations such as rotations, flips, elastic deformations, contrast optimization, and noise modulation. For the final evaluation, we ensembled the best-performing models from 5-fold validation using majority voting to generate consensus segmentation masks.

### Post-processing and CTAC Classification

The U-Net output was refined through a structured post-processing pipeline that included probability thresholding, small artifacts removal, morphological smoothening, and watershed-based instance segmentation to separate adjacent cells. Classification was performed in two integrated stages combining U-Net segmentation with a random forest model. In the first stage (Feature Extraction), a total of 64 features were computed for each segmented candidate: morphological (25, e.g., area, perimeter, solidity, eccentricity, axis lengths, and convexity), intensity-based (18, mean/median/max intensity, contrast, and entropy in EPCAM/Hoechst 33342 channels), texture-related (13, Haralick descriptors such as energy, correlation, and homogeneity) and spatial context (8: proximal intercellular distances, and local density)[24,25]. In the second stage (Classification), the 64-dimensional feature vector feeds into a random forest classifier (100 trees, max depth = 10, min samples per leaf = 5) using the entropy criterion and class weights that are inversely proportional to class frequencies[26,27,28,29]. CTACs were identified based on defined criteria including cell diameter (>10 μm), EPCAM signal-to-background ratio (>2.5), nuclear-to-cytoplasmic ratio (>0.7), and characteristic epithelial morphology.

### Analytical Validation

To ensure suitability for clinical use, we conducted comprehensive analytical validation of the qualitative CTAC detection model. The validation protocol assessed critical performance parameters including the limit of detection, analytical specificity against commonly encountered non-malignant cells, precision (repeatability and reproducibility), sample stability, and potential interference from exogenous substances. These validation studies were aimed to establish the analytical performance characteristics required for clinical implementation. Detailed protocols are provided in the supplementary materials.

### Exploratory Study

The objective of the study was to demonstrate that CTACs are detected in advanced cancers and that CTACs are undetectable (or rarely detected) in asymptomatic individuals, i.e., the sensitivity and specificity of the approach. This study included peripheral blood specimens from asymptomatic individuals (n = 428) and patients (n = 354) with advanced, recurrent solid tumors. Demographics of the sub-cohorts are provided in Table 1. All blood specimens were processed as per the established workflow to determine the detection of CTACs. The Proof of Concept (PoC) study sub-cohorts were stratified into training, validation and test sets and evaluated as described in a previous section. Figure 4 depicts the various clinical study (sub-)cohorts.

**Figure 1:**
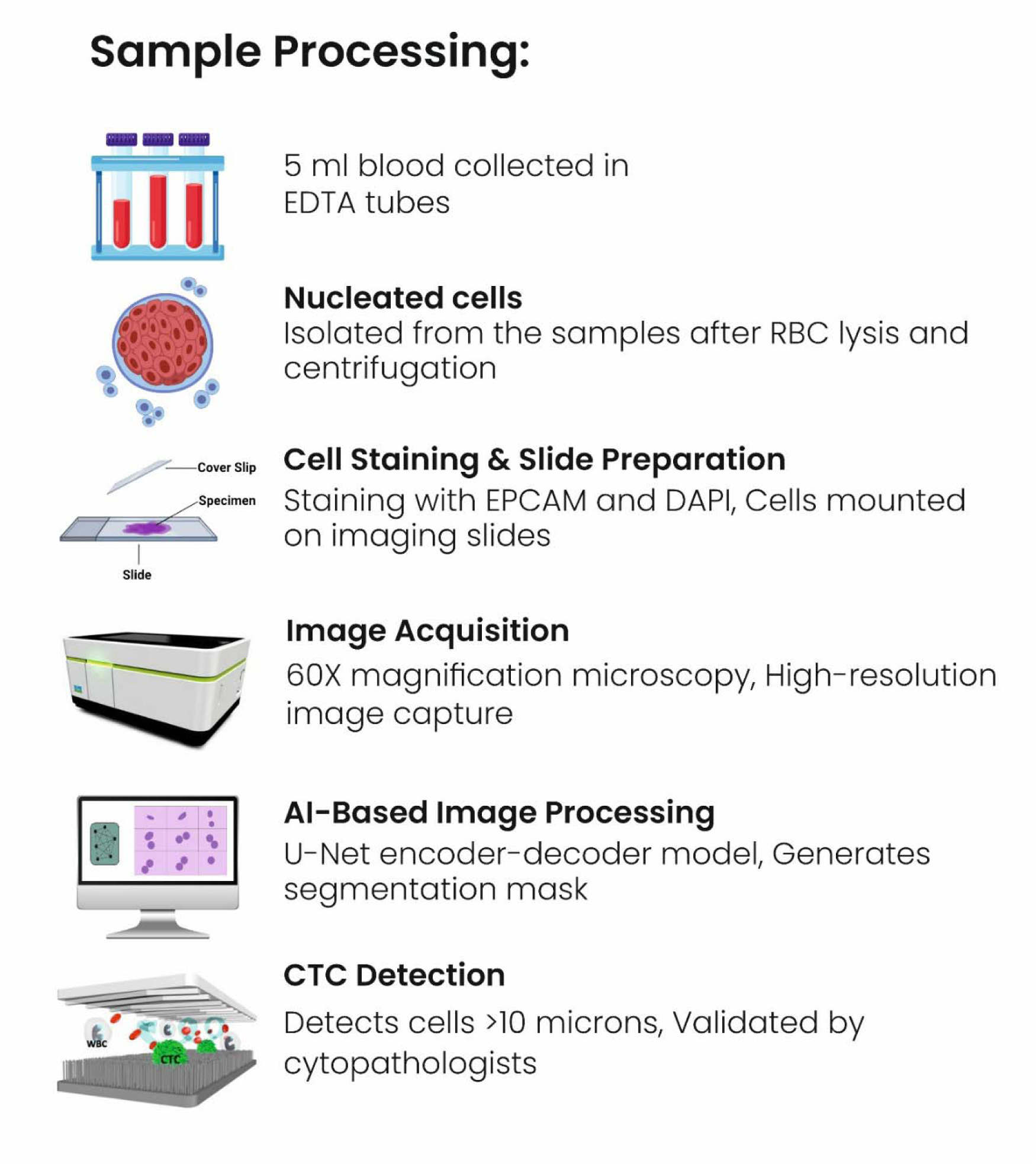
Sample processing workflow diagram showing the steps from blood collection to slide preparation for imaging

**Figure 2:**
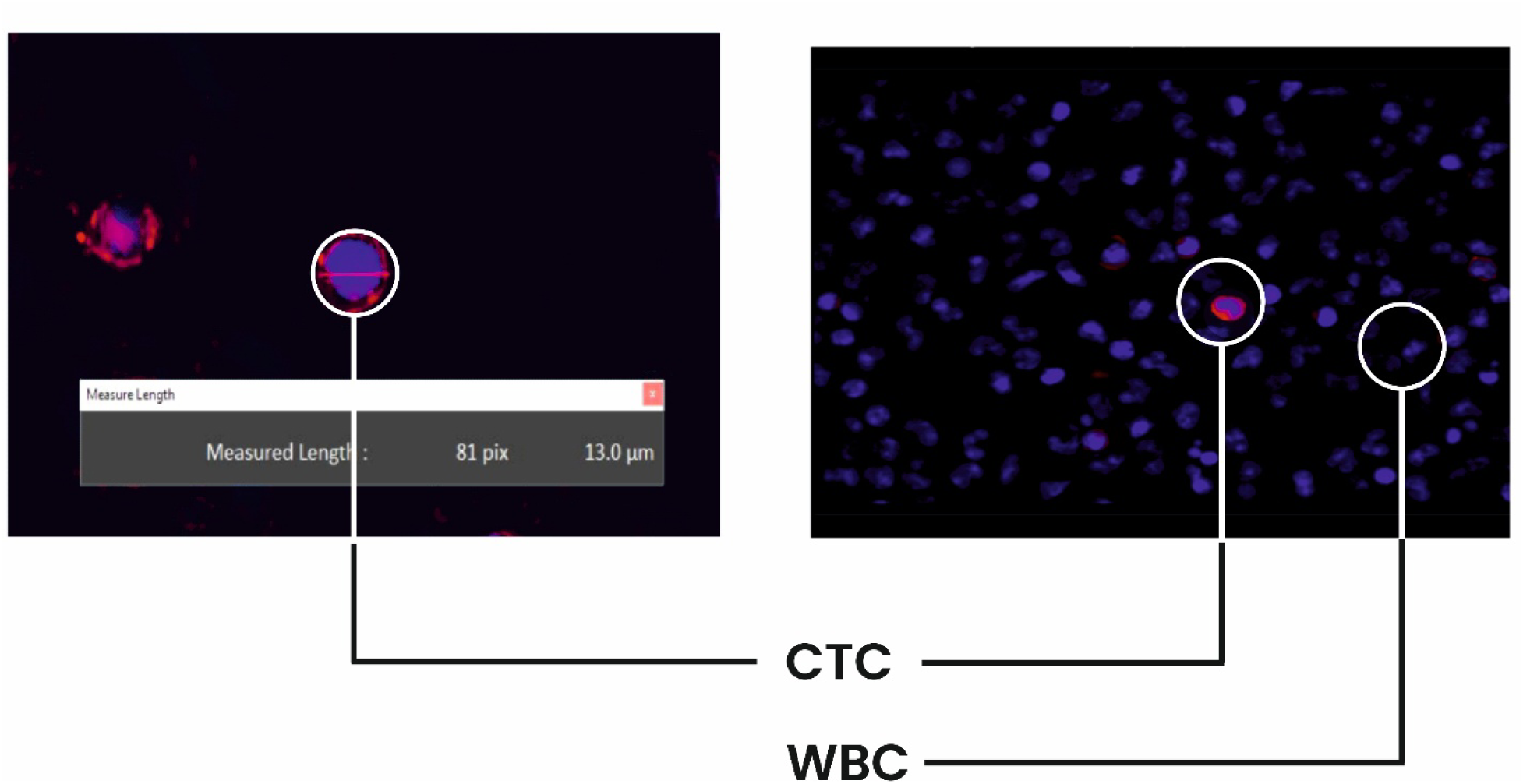
Representative images showing annotation of CTACs versus non-CTACs, highlighting the morphological and fluorescence characteristics used for discrimination

**Figure 3:**
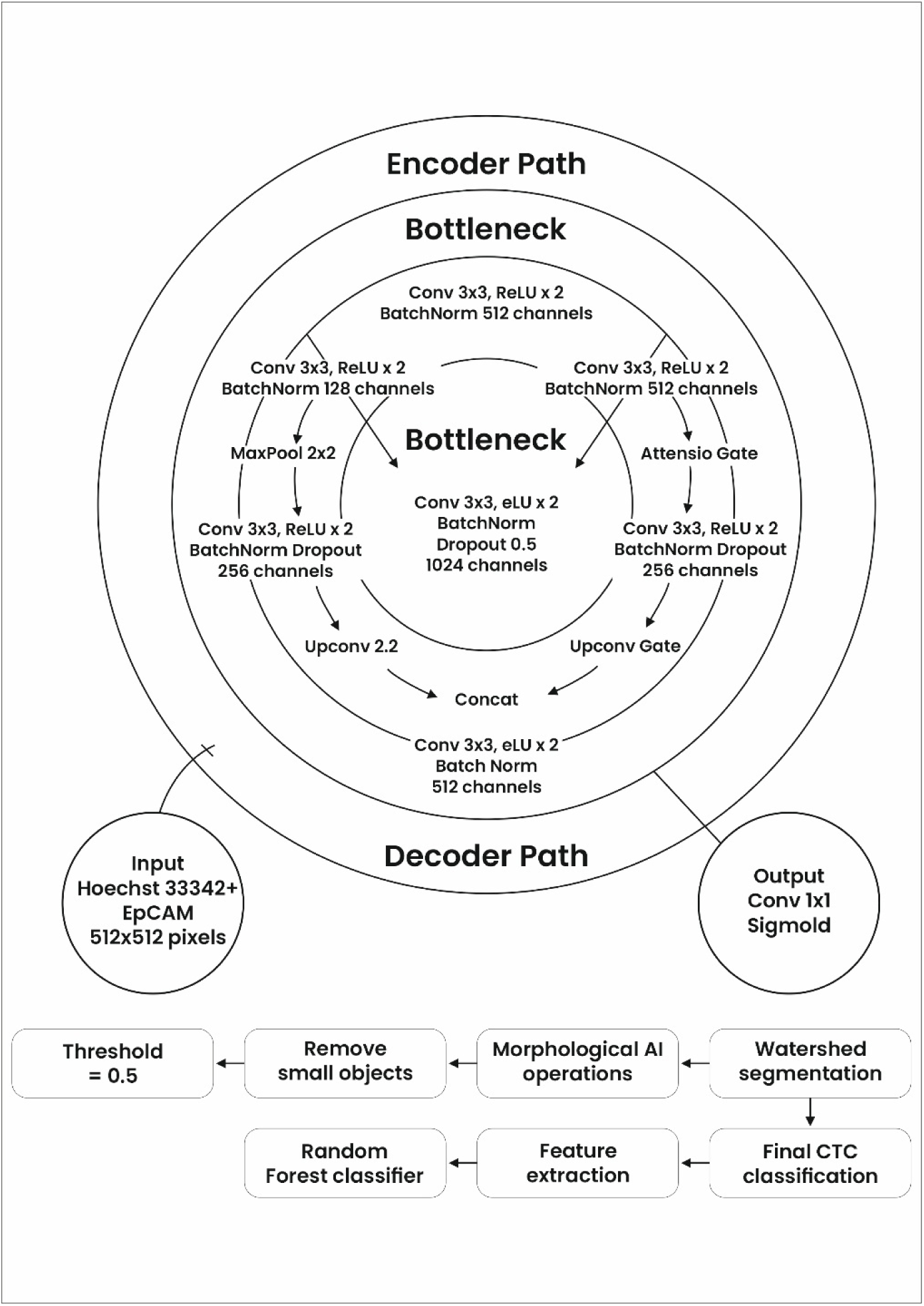
Attention-Enhanced U-Net Architecture and Post-Processing Workflow for Circulating Tumor Associated Cell (CTAC) Detection. Overview of the attention-enhanced U-Net pipeline for CTAC detection from dual-channel (Hoechst/EpCAM) fluorescence images, showing the encoder–decoder architecture with skip connections and attention gates, followed by post-processing, feature extraction, and classification to generate final CTAC calls.

**Figure 4:**
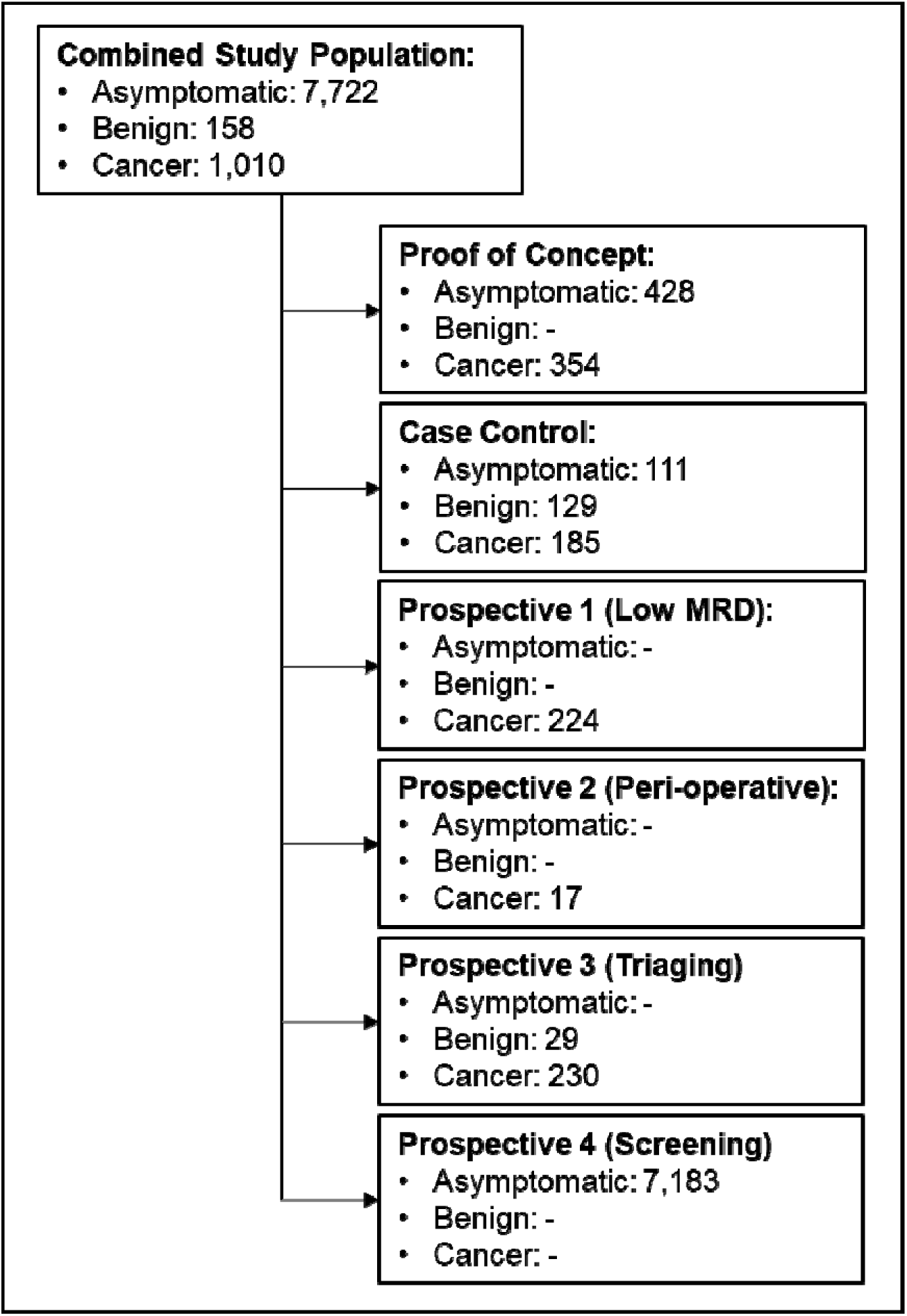
Clinical Study Cohorts. Diagrammatic representation of the overall study population as well as sub-cohorts.

**Table 1.** Study-wise demographics of patients with cancer and non-malignant (benign) conditions.

| **Study** | **Proof of Concept** | **Case Control** |  | **Low Tumor Burden** | **Peri-Operative** | **Suspected Cases (Final Diagnosis)** |  |
| --- | --- | --- | --- | --- | --- | --- | --- |
| **Type** | **Cancer** | **Cancer** | **Benign** | **Cancer** | **Cancer** | **Cancer** | **Benign** |
| **n =** | **354** | **185** | **129** | **224** | **17** | **230** | **29** |
| **Gender** |  |  |  |  |  |  |  |
| **Male** | 115 | 88 | 83 | 79 | 11 | 80 | 11 |
| **Female** | 239 | 97 | 46 | 145 | 6 | 150 | 18 |
| **Age (years)** |  |  |  |  |  |  |  |
| **Median** | 57 | 55 | 60 | 64 | 52 | 53 | 50 |
| **Range** | 24-88 | 20-83 | 17-88 | 21-88 | 26-76 | 19-92 | 17-77 |
| **Organ** |  |  |  |  |  |  |  |
| Breast | 98 | 51 | 8 | 54 | 1 | 82 | 8 |
| Colorectal | 83 | 22 | - | 56 | 5 | 12 | - |
| Lung | 26 | 1 | 4 | 23 | - | 23 | 4 |
| Ovary | 34 | 7 | 16 | 21 | - | 24 | 6 |
| Pancreas | 26 | 6 | 12 | 18 | - | 11 | - |
| Prostate | 9 | 9 | 46 | 5 | - | 14 | 6 |
| Head and Neck | 6 | 57 | 2 | 2 | 7 | 33 | 2 |
| Esophagus | 9 | 1 | - | 5 | - | 1 | - |
| Uterine | 12 | 7 | - | 6 | 1 | 3 | - |

| Study | Proof of Concept | Case Control |  | Low Tumor Burden | Peri-Operative | Suspected Cases (Final Diagnosis) |  |
| --- | --- | --- | --- | --- | --- | --- | --- |
| Gastric | 12 | 5 | - | 7 | - | 1 | - |
| Unknown | 10 | 1 | 31* | 3 | - | 3 | - |
| Kidney | 5 | 3 | 1 | 4 | - | 4 | - |
| Hepatobiliary | 12 | - | 3 | 7 | - | 4 | 2 |
| Bladder | 2 | - | - | 3 | - | 3 | - |
| Cervix | 1 | 4 | 1 | - | 2 | 10 | 1 |
| Thyroid | 4 | 5 | 5 | 3 | - | 1 | - |
| Thymus | 2 | - | - | 2 | - | 1 | - |
| Anal | 2 | - | - | 2 | - | - | - |
| Penis | 1 | 3 | - | 1 | - | - | - |
| Small bowel | - | 1 | - | 2 | - | - | - |
| Vulva | - | 2 | - | - | 1 | - | - |

*\*includes conditions such as stroke, diabetes, hypertension, pyrexia or infection*

#### Clinical Studies

The clinical performance of the model to detect cancer cases (sensitivity) and differentiate asymptomatic individuals or those with benign conditions (specificity) was evaluated in one case-control and four prospective clinical studies. All studies were performed independently. All studies used peripheral blood collected at baseline or leftover specimens. There were no follow-up specimens collected in any study.

##### Case Control Study

The study cohort (Table 1, Table 2) included stage I/II therapy naive cancer patients (n = 185), patients with benign conditions (n = 129), and asymptomatic volunteers with no history of cancer diagnosis (n = 111). With 185 cancer cases and 240 non-cancer controls, the study size allowed overall performance to be estimated with single-digit percentage precision. For a 90% estimated sensitivity, the 95% CI would be (85.7%, 94.3%), and for a 95% estimated specificity, the 95% CI would be (92.2%, 97.8%), both indicating acceptable sensitivity and specificity estimation.

**Table 2.** Study-wise demographics of asymptomatic individuals.

| **Study** | **Proof of Concept** | **Case Control** | **Cancer Screening** |
| --- | --- | --- | --- |
| **n =** | **428** | **111** | **7183** |
| **Gender** |  |  |  |
| **Male** | 214 | 63 | 3218 |
| **Female** | 214 | 48 | 3965 |
| **Age (years)** |  |  |  |
| **Median** | 46 | 44 | 46 |
| **Range** | 20-83 | 20-94 | 20-94 |

##### Prospective Study 1 (Low Radiological Tumor Disease)

The study cohort (Table 1) included 224 patients with various solid tumors showing objective (complete or significant) response to anticancer treatments. The study determined the sensitivity of the model, i.e., the ability to detect tumors with lower burden.

##### Prospective Study 2 (Perioperative CTAC Dynamics)

This study evaluated peri-operative CTAC detectability in patients with solid tumors (Table 1, Supplementary Table 2) to establish a qualitative link between CTACs and tumor by demonstrating that the abrogation of the cause (tumor) impacts the effect (CTAC detection). In this study, each patient was their own control. For this study, pre-surgery (-4 h) and post-surgery (+24 h) blood specimens were collected from recently diagnosed, systemic therapy naive patients (n = 17) with resectable solid tumors and evaluated.

##### Prospective Study 3 (Cancer versus Benign Specificity)

The study cohort (Table 1) included 259 individuals suspected of various cancers based on clinical findings or symptoms. All patients underwent standard-of-care (SoC) tissue-based histopathological diagnosis. The objective of the study was to determine the ability of the model to support diagnostic triaging in suspected cancer cases.

##### Prospective Study 4 (MCED in Cancer Screening Population)

The study cohort (Table 2) included 7183 asymptomatic individuals; 3218 males and 3965 females with a median age of 46 years (range: 20 - 94 years) who had availed of the sponsor’s CTAC based multi-cancer early detection (MCED) test. Leftover specimens were processed as per the described method. Individuals with CTAC positive findings (as per the approach described in this manuscript) were clinically followed up to determine cancer diagnosis. The objective of this study was to evaluate the suitability of the model for MCED in an unselected cancer screening population.

## Results

### Analytical Validation

Findings of the analytical validation studies are provided in the supplementary materials.

### Exploratory Study

In this study, the model achieved an overall 90.68% sensitivity (95% CI: 87.16%, 93.50%) for CTAC-detection in advanced cancers (Table 3). The sensitivity ranged from 66.67% (Head and Neck cancers) to 100% (Gastric, Uterine, Esophageal, Prostate, Kidney, Hepatobiliary). CTACs were undetectable in 99.53% (95% CI: 98.32%, 99.94%) asymptomatic individuals with no history of cancer diagnosis. The study findings established acceptable sensitivity and specificity of the approach.

**Table 3.** CTAC Detection in Cancers. Consolidated CTAC detection rates observed in various cancer cases across all clinical study cohorts. Where the number of any cancer-type in any study is <5, the findings are grouped under ‘Others’

| Cancer Type | CTAC Positive Rates (n/N, %) in Cancers in Various Study Cohorts |  |  |  |
| --- | --- | --- | --- | --- |
|  | Proof of Concept | Case Control | Low-Tumor Burden | Prospective |
| Breast | 88/98 (89.80%) | 44/51 (86.27%) | 50/54 (92.59%) | 75/82 (91.46%) |
| Colorectal | 72/83 (86.75%) | 21/22 (95.45%) | 49/56 (87.50%) | 9/12 (75.00%) |
| Ovary | 32/34 (94.12%) | 6/7 (85.71%) | 19/21 (90.48%) | 17/24 (70.83%) |
| Lung | 24/26 (92.31%) | - | 21/23 (91.30%) | 17/23 (73.91%) |
| Pancreas | 24/26 (92.31%) | 5/6 (83.33%) | 18/18 (100%) | 9/11 (81.82%) |
| Gastric | 12/12 (100%) | - | 7/7 (100%) | - |
| Uterine | 12/12 (100%) | 6/7 (85.71%) | 6/6 (100%) | - |
| Unknown primary | 8/10 (80.00%) | - | - | - |
| Esophagus | 9/9 (100%) | - | 5/5 (100%) | - |
| Prostate | 9/9 (100%) | 9/9 (100%) | 5/5 (100%) | 13/14 (92.86%) |
| Hepatobiliary | 12/12 (100%) | - | 5/5 (100%) | - |
| Head and neck | 4/6 (66.67%) | 48/57 (84.21%) | - | 21/33 (63.64%) |
| Kidney | 5/5 (100%) | - | - | - |
| Cervix | - | - | - | 8/10 (80.00%) |
| Others | 10/12 (83.33%) | 25/26 (96.15%) | 21/24 (87.5%) | 14/21 (66.67%) |
| **Total** | **321/354 (90.68%)** | **164/185 (88.65%)** | **206/224 (91.96%)** | **184/230 (80.00%)** |

#### Clinical Studies

##### Case Control Study

In the case-control study, the model demonstrated 88.65% sensitivity (95% CI: 83.17%, 92.83%) for detecting Stage I/II cancers (Table 3). The sensitivity ranged from 83.33% (Pancreatic cancer) to 100% (Prostate cancer). In this study also, lower CTAC-detection rate was observed for Head and Neck cancer (84.21%). In this cohort, CTAC detection rate appeared to be inversely proportional to the grade, with 75% sensitivity in Low Grade (well-differentiated, n = 20), 87.93% sensitivity in Moderate Grade (moderately differentiated, n = 51) and 97.5% sensitivity in High Grade (poorly or un-differentiated, n = 40) tumors. There were no significant differences in CTAC detection rates based on gender (87.5% in males vs. 89.9% in females). Among the individuals with benign conditions, the model had 78.95% specificity (95% CI: 71.03%, 85.53%) with CTACs being undetectable in 105 of 133 cases. CTACs were detected in 15/46 (32.61%) cases of benign prostate conditions, suggesting undiagnosed or latent malignancy; while the need for follow-up has been communicated to the oncologist, no follow-up data is available at the time of submission. CTACs were undetectable in all specimens from asymptomatic individuals, yielding 100% specificity (95% CI: 96.73%, 100%). The findings of the case-control study suggest robust clinical performance characteristics that support the intended clinical utility of the model.

##### Prospective Study 1 (Low Tumor Burden)

The first prospective cohort included diagnosed and pretreated solid tumor cases (n = 224) with complete or significant treatment response (post SoC). The objective of this study was to determine the utility of the model for non-invasive post-treatment monitoring of disease status in patients with radiologically low or undetectable tumor burden. In this cohort of patients, the model achieved 91.96% CTAC-detection sensitivity (95% CI: 87.60%, 95.17%). CTAC detection sensitivity ranged from 87.5% (Colorectal) to 100% (Pancreaticobiliary, Gastroesophageal, Uterine, Prostate). The detection sensitivity was comparable in females (93.10%, n = 145) and males (91.14%, n = 79). (Table 3).

##### Prospective Study 2 (Perioperative CTAC Dynamics)

In the perioperative study, CTACs were detected in all (n = 17) pre-surgery blood specimens. In the post-surgery specimens CTACs were detectable in 5 cases (29.41%) and undetectable in 12 cases (70.59%). The study findings support a causal relationship between qualitative tumor burden and CTAC detectability.

##### Prospective Study 3 (Cancer versus Benign Specificity)

This prospective cohort included patients (n = 259) with no prior diagnosis of cancer, who presented with clinical symptoms, clinical laboratory findings, or radiological findings suspected of cancer necessitating the standard of care (SoC) diagnostic pathway based on histopathological examination (HPE) of tissue sample. In this cohort, 191 patients were CTAC-positive and 68 patients were CTAC-negative. Of the 259 suspected cases, histopathological diagnosis confirmed cancer in 230 cases and indicated benign conditions in 29 cases. When the CTAC detection status was correlated with histopathological diagnosis, the model demonstrated 96.34% PPV (95% CI: 93.22%, 98.05%) and 32.35% NPV (95% CI: 25.58%, 39.95%), based on a sensitivity of 80.00% (95% CI: 74.24%, 84.97%) and a specificity of 75.86% (95% CI: 56.46%, 89.70%) (Table 3). When adjusted for a ∼25% cancer diagnosis rate among suspected cases the PPV and NPV were 52.49% (95% CI: 36.61%, 67.88) and 91.92% (95% CI: 89.11%, 94.06%), respectively.

##### Prospective Study 4 (MCED in Cancer Screening Population)

Among the 7183 asymptomatic individuals, 7139 (99.39%) were CTAC negative and 44(0.61%) were CTAC positive. Among the 7139 CTAC negative cases, 2 (0.03%) were diagnosed with Stage 0 / I cancers. Among the 44 CTAC positive cases, 16 (36.4%) were diagnosed with Stage I / II cancers including prostate (n=6), breast (n=3), colon(n=2), cervix (n=2), stomach (n=2), or esophagus (n=1). In 10 CTAC positive cases, there was no radiologically detectable evidence of disease. Since follow-up was ongoing for the remaining 18 CTAC positive cases at the time of submission, they were provisionally considered as false positives. In this cohort, the PPV and NPV estimates were 36.36% (95% CI: 27.61%, 46.12%) and 99.97% (95% CI: 99.90%, 99.99%), based on a provisional specificity of 99.61% (95% CI: 99.44%, 99.74%). When adjusted for 0.1% cancer prevalence among asymptomatic individuals, the PPV and NPV estimates were 18.55% (95% CI: 13.19%, 25.43%) and 99.99% (95% CI: 99.96%, 100%), respectively. These PPV and NPV estimates are initial and may improve upon further follow-up.

## Discussion

This study demonstrates the efficacy of a customized U-Net architecture with attention-enhanced skip connections for automated CTAC detection from peripheral blood samples of individuals with early, advanced or recurring malignancies. The strong performance characteristics in the case-control and prospective cohorts are relevant for disease monitoring after curative intent and MCED. Our approach enhances the accuracy and objectivity of CTAC detection, and could significantly impact cancer management strategies, by enabling earlier and more personalized clinical decisions.

Conventional CTC assays have been constrained by low sensitivity, low specificity, lack of harmonized standards for interpretation, and inadequately established clinical utility value or actionability, all of which may have contributed to a gradual decline in enthusiasm for CTCs as robust clinical biomarkers [3,4]. The historical limitations that may have undermined the reliability of CTC-based technologies arise from methodological and analytical constraints and not from the biological characteristics of CTCs themselves [3,30]. Our framework leveraged pixel-level segmentation (rather than bounding box-based detection) to achieve precise characterization of cellular and nuclear morphology based on biomarker localization (fluorescence signal) patterns. It also incorporated attention mechanisms into skip connections to enable (selective) emphasis on discriminative features, and suppress background noise, thereby improving CTAC detection within a significantly larger PBNC population. The consistent performance across diverse cancer types suggests that the model captured generalizable morphological and fluorescence (biomarker positivity and localization) features of CTACs and was not prone to confounding effects of tumor cancer-specific variabilities. Our model uses predefined size exclusion and nuclear-to-cytoplasmic (N:C) ratio parameters in conjunction with biomarker (EpCAM) positivity for CTAC detection. While EpCAM status provides molecular specificity to confirm epithelial tumor (but not the organ of origin), the biophysical and morphological filters provide orthogonal verification to improve detection accuracy[31]. Together, they reduce false positives from large or atypical leukocytes, and yield a more robust and reliable identification of CTACs across heterogeneous tumor states. This model addresses several key analytical limitations that have undermined other approaches. The findings demonstrate that meaningful technological refinement can yield objective gains in accuracy and reproducibility, which reaffirm the credibility of CTACs/CTCs as a reliable biomarker in oncology.

The applicability of the model was evaluated in case-control as well as in prospective studies. Robust performance characteristics in all specimens (controlled study and real-world), suggests that analytical and biological variability in field samples does not significantly degrade model accuracy. These strengths support its potential pan-solid tumor clinical utility.

The peri-operative study findings demonstrate the specificity of CTAC detection via direct association with tumor burden, and support the clinical relevance of the method. The findings support serial CTAC monitoring as a surrogate marker of therapeutic response (or early recurrence), complementing radiologic or serologic follow-up. Further evaluation in multi-center, ethnically diverse populations will be critical to confirm this generalizability.

The integration of attention-enhanced skip connections represents a significant advancement over the original U-Net architecture[16], by preventing the propagation of irrelevant spatial information, allowing the model to focus on salient (discriminative) cellular characteristics. Combining U-Net segmentation with random forest classification of extracted morphological, intensity, and texture features added interpretability and robustness, reducing misclassification of non-CTAC artifacts. Our hybrid architecture for interpretation of extracted feature sets (that enables biological correlation) supports more informed clinical interpretation and decision making as compared to conventional ‘black-box’ deep learning classifiers. This approach improves the automated detection of CTACs especially against a background of non-malignant blood cells reducing the dependence on laborious manual assessments as well as instances of user-fatigue and fatigue induced errors.

The U-Net’s encoder-decoder structure is well-matched to the detection problem. The encoder progressively compresses spatial information through convolutional and pooling layers, building increasingly abstract feature representations at each level – from fine details such as membrane-localised EpCAM fluorescence and nuclear texture at shallow layers, to broader cellular geometry and size relationships at deeper layers. The decoder reconstructs this back to full resolution for precise cell boundary delineation. This multi-scale hierarchy maps naturally to the definition of CTACs: their identification rests on features at different spatial scales, from EpCAM signal at the membrane, through nuclear morphology, to the N:C ratio. A single-scale classifier would trade off between fine detail and broader context; the U-Net avoids this by maintaining representations at every scale through the skip connections, while the attention gates learn which scale is most informative at each location. The model can therefore preserve discrimination even when no single feature alone would suffice to separate a CTAC from an atypical leukocyte or debris.

The performance characteristics of the CTAC detection framework support multiple clinical applications, including early detection of common epithelial cancers, diagnostic triaging, assessment of treatment response, and MRD monitoring[32]. The high PPV (irrespective of lower NPV) in the cohort of suspected cancer cases, supports its potential clinical application as a rule-in (triaging) test to identify those who are more likely to benefit from further follow-up investigations. On the other hand, the high negative predictive value (NPV) in asymptomatic populations supports its application as an effective rule-out test to reduce unnecessary intensive / invasive procedures in low(er)-risk individuals [33,34,35,36]. The reported PPV and NPV in the asymptomatic (screening) cohort are initial estimates that may improve upon availability of further / long-term follow up data. Despite this limitation, the PPV and NPV estimates support the potential of test findings to support more effective clinical decision making within the standard of care (SoC) framework. Illustratively, individuals with CTAC-negative status may be encouraged to continue participation in standard of care (SoC) cancer screening, whereas those with CTAC-positive status may be considered for additional evaluations or follow-up. This approach integrates into existing healthcare pathways and may help minimize delays in time to diagnosis. As a non-invasive (and non-intrusive) liquid biopsy test, the approach may even help improve compliance with cancer screening.

In addition to the perceived benefits, we acknowledge the following possible limitations. The reliance on EpCAM as the sole epithelial marker may overlook CTACs with low or no epithelial features (e.g., mesenchymal phenotypes), potentially impacting detection (sensitivity). While this has been a limitation of EpCAM-based CTAC immuno-enrichment methods, EpCAM immuno-fluorescence staining has higher sensitivity for CTAC detection. We intend to evaluate additional biomarkers to improve CTAC detection and reduce residual false negative risks. The present study focusses on epithelial malignancies, and its utility or performance in non-epithelial cancers is not yet established. The case-control clinical study design may overestimate real-world performance compared to a prospective screening context. However, this is partially mitigated by consistent results from the inclusion of unselected populations. The PPV observed in the cancer screening cohort (n = 7183) reflects the rarity of early-stage cancers in asymptomatic populations and highlights the importance of follow-up confirmation using imaging or other diagnostic modalities. The incomplete longitudinal outcome data for the screening cohort participants limits definitive assessment of long-term clinical benefit and interval cancer incidence. Tumor burden was not operationalized using rigid quantitative thresholds, and classification into low- or high-burden categories therefore relied on clinical (or radiological) interpretation. The model is currently intended for qualitative CTAC detection, and not quantitatively correlate CTACs with tumor volume or disease kinetics. The model is currently not intended to provide information on the organ or tissue of origin (cancer type and subtype) in any application, including MCED. The primary objective of MCED is to identify biologically significant malignancy at an early stage to improve clinical outcomes. Hence a more appropriate metric for evaluating MCED would be sensitivity for Stage I/II disease. Tissue localization is clinically informative but contingent on successful detection and depends on imaging and histopathological confirmation. Methods that rely predominantly on tumor-derived molecular shedding, including methylation-based assays, may have lower performance in early-stage and low-shedding cancers, where analyte abundance is constrained. In contrast, detection of systemic signals of malignancy could potentially enable timely diagnostic escalation even when anatomical attribution is indeterminate. This hierarchy preference of detection over localization aligns with real-world expectations and oncology workflows.

Future, statistically powered, prospective, longitudinal studies shall validate this approach for screening, MRD and therapeutic monitoring, in a wider scope of epithelial cancers and in ethnically diverse populations. The CTAC-based MCED screening has the potential to become an affordable liquid biopsy modality for population level screening. Despite these limitations, the study findings demonstrate that a layered, attention-enhanced deep learning approach can robustly detect rare CTACs, preserve interpretability, and thus re-establish the viability of CTAC-based MCED. Our study findings support the case for the clinical utility of AI-integrated cytological diagnostics.

## Conclusions

The enhanced U-Net model has compelling advantages over conventional convolutional neural network (CNN) classifiers, where spatial information may degrade during down-sampling. Our model demonstrated robust performance in early-stage (and low burden) cancers, with consistent detection rates and medically relevant correlations in unselected real-world populations. The findings represent advancements in liquid biopsy technology with promising clinical practice implications in cancer screening (especially MCED), diagnostic triaging, and monitoring.

## Abbreviations

AI: Artificial Intelligence
CI: Confidence Interval
CDX2: Caudal Type Homeobox 2, colon marker
CNN: Convolutional Neural Network
CTAC: Circulating Tumor Cell
DCIS: Ductal Carcinoma In Situ
EpCAM: Epithelial Cell Adhesion Molecule
FOV: Field of View
GAN: Generative Adversarial Network
GATA3: Transcription factor, breast marker
H&E: Hematoxylin and Eosin (staining)
ICC: Immunocytochemistry
MCED: Multi-Cancer Early Detection
NC: Negative Control
NKX3.1: Homeobox gene, prostate marker
NPV: Negative Predictive Value
PAX8: Paired Box Gene 8, ovary marker
PBNC: Peripheral Blood Nucleated Cell
PBS: Phosphate-Buffered Saline
PC: Positive Control
PPV: Positive Predictive Value
RBC: red blood cell
ReLU: Rectified Linear Unit
SOX9: SRY-Box Transcription Factor 9, pancreas marker
STR: Short Tandem Repeat
TTF1: Thyroid Transcription Factor 1, lung marker

## Acknowledgments

The authors express gratitude to all human volunteers who provided biological specimens (peripheral blood) for the purpose of this study. The authors also acknowledge patients and volunteers who consented to the use of their deidentified leftover specimens for research and scientific purposes. The authors acknowledge the contributions of Mr. Chetan Patil and Mr. Akash Prashad towards study conduct.

## Author Contributions

Conceptualization: MC, TC, AG, RP, AdS, SL, NR, HAS, PA, SS, RD;

Data Curation: SP, PS, SG;

Formal Analysis: RD, SP, PS, SG;

Funding Acquisition: RD, VD, DP;

Investigation: RD, SP, PS, SG, AP, CP;

Methodology: MC, TC, AG, RP, AdS, SL, NR, HAS, PA, SS;

Project Administration: RD, VD, DP;

Resources: RD, VD, DP;

Software: RD;

Supervision: RD, SP,

Validation: RD, SP;

Visualization: MC, TC, AG, RP, AdS, SL, NR, HAS, PA, SS;

Writing - Original Draft Preparation: RD, AS;

Writing - Review & Editing: MC, TC, AG, RP, AdS, SL, NR, HAS, PA, SS;

## Conflict of Interest

HAS and AdS report no conflicts of interest associated with the scope of the present study. MC, SL, NR, TC, AG and RP declare fractional stock ownership of the study sponsor. DP, VD, DA, SS, PA, SP, SA, PS, SG, AjS are employees of the study sponsor. RD is the founder of the study sponsor.

## Ethics Statement

All studies with human specimens were conducted in compliance with Ethical Practices, Good Clinical Practices and in accordance with the Declaration of Helsinki. Informed Consent was obtained from all human volunteers prior to specimen collection, or for the use of leftover specimens for research purposes and publication of deidentified data and findings. None of the human volunteers underwent any invasive procedure solely for the purpose of this study, and any invasive procedures mentioned in this manuscript were standard of care (SoC) cancer management procedures.

## Data Availability

More comprehensive descriptions of the U-Net structure and the details on hyperparameter training can be made available on request under appropriate non-disclosure agreements.

## Notes

### Funding Statement

This study did not receive any funding

### Author Declarations

Institutional Ethics Committee (IEC) of the Study Sponsor (Datar Cancer Genetics, DCG) gave ethical approval for this work.

