## Supplementary Materials for "Attention-Enhanced U-Net Segmentation for Reliable Detection of Circulating Tumor-Associated Cells"

**Summary of Analytical Validation**

The analytical performance characteristics of the process was evaluated in structured studies.

**Accuracy:** Accuracy was established on contrived specimens generated using MCF7, breast cancer cell line that shows consistent high EpCAM positivity. Peripheral blood collected from healthy donors (HD; n = 15; 9 male, 6 female; median age 38 years, range 24-56 years) was used for this study. Fifteen 1 mL aliquots 1 each from all 15 individuals) were unspiked, fifteen 1 mL aliquots (1 each from all 15 individuals) were spiked with 100 MCF7 cells, and five 1 mL aliquots (1 each from 5 individuals) were spiked with 50 MCF7 cells. All samples were processed as per the test workflow for detection of EpCAM+ cells. The prespecified acceptance criteria were >90% positive percent agreement (PPA) and >95% negative percent agreement (NPA). EpCAM+ cells were detected in all spiked specimens (n = 20), while no marker-positive cells were detected in unspiked specimens (n = 15). Based on this, the test had a PPA of 100% (95% CI: 83.16%, 100.00%), NPA of 100% (95% CI: 78.20%, 100.00%), and overall agreement of 100% (95% CI: 90.00%, 100.00%).

**Precision:** Precision was established using contrived specimens. generated using MCF7 (breast cancer) cell line that shows consistent high EpCAM positivity. HD peripheral blood pool was used for this study. HD blood samples (5 mL) were spiked with MCF7 cells to generate positive samples, while unspiked HD blood aliquots (5 mL) were used as negative samples. All samples were processed as per the test workflow for detection of EpCAM+ cells. A total of 77 positive samples and 77 negative samples were processed by 2 operators, over >20 days on a single instrument. Precision was established from the PPA and NPA in positive and negative samples respectively. The prespecified acceptance criteria were >95% PPA and >99% NPA. EpCAM+ cells were detected in all 77 positive samples yielding a PPA of 100% (95% CI: 95.32%, 100%). EpCAM+ cells were undetectable in all 77 negative samples yielding a NPA of 100% (95% CI: 95.32%, 100%).

**Specificity:** The Specificity of the test was established using contrived specimens. generated using SJCRH30, U87MG, and A375 cells that show consistent negative status for EpCAM. HD peripheral blood pool was used for this study. HD blood samples (5 mL) were spiked with 5, 10, 50, 100, and 200 of each type of cells (multiple replicates). All samples were processed as per the test workflow for detection of EpCAM+ cells. Specificity was established from the NPA. The prespecified acceptance criteria was >99% NPA. EpCAM+ cells were undetectable in all samples yielding 100% NPA.

**Limit of Blank (LoB):** The Limit of blank of the test was established using HD blood samples from 10 volunteers (5 male, 5 female; age range 39-56 years) with no prior diagnosis of cancer and no current suspicion of cancer. 5 mL blood samples were collected and processed as per the test workflow for detection of EpCAM+ cells. The Limit of Blank for qualitative determination was established from the NPA in the known negative samples. The prespecified acceptance criteria was >99% NPA. EpCAM+ cells were undetectable in all negative samples yielding NPA of 100% (95% CI: 78.20%, 100.00%).

**Limit of Detection (LoD):** The Limit of detection of the test was established using contrived specimens. generated using MCF7 (Breast Cancer), SW620 (Colon Cancer), PL45 (Pancreatic Cancer), OVCAR3 (Ovarian Cancer), PC3 (Prostate Cancer) cell lines that show consistent high EpCAM positivity. HD peripheral blood pool was used for this study. HD blood samples (5 mL) were spiked with each type of cell lines at 1, 2, 5, 10, 50 and 100 cells with multiple replicates per spike level. All samples were processed as per the test workflow for detection of EpCAM+ cells. The LoD was determined as the lowest number of spiked cells that could be detected in at least 50% of the replicates. Based on these criteria, the LoD was determined to be 2 cells / mL.

The complete analytical validation datasets may be obtained from the authors upon request.

**Supplementary Tables.**

**Supplementary Table 1. Demographics of the Proof-of-Concept Study Cohorts.**

| **Cancer type** | **Plausibility Study** | **Causality Study** |
| --- | --- | --- |
| **Gender**  **Male**  **Female** | 886  1440 | 14  9 |
| **Age (years)**  **Median**  **Range** | 59  20-90 | 63  26-76 |
| **Cancer Types** |  |  |
| Breast | 685 | 3 |
| Colorectal | 357 | 7 |
| Lung | 266 | - |
| Ovary + Peritoneum | 171 | - |
| Pancreas | 159 | - |
| Prostate | 115 | - |
| Head and Neck | 109 | 8 |
| Esophagus | 76 | - |
| Uterine | 61 | 1 |
| Gastric | 54 | - |
| Unknown Primary | 42 | - |
| Kidney | 40 | - |
| Bile Duct + Gallbladder | 55 | - |
| Bladder | 35 | - |
| Cervix | 25 | 2 |
| Thyroid | 21 | - |
| Liver | 12 | - |
| Testes | 7 | - |
| Thymus | 8 | - |
| Anal | 6 | - |
| Adrenal | 5 | - |
| Penis | 3 | - |
| Small intestine | 3 | - |
| Vulva | 3 | 1 |
| Basal cell carcinoma | 1 | - |
| **Grand Total** | **2319** | **22** |

**Supplementary Table 2. Demographics of the Case-Control Study Cohorts.**

| **Cohorts** | **Cancer** | **Benign** | **Asymptomatic** |
| --- | --- | --- | --- |
| **Gender**  **Male**  **Female** | 88  97 | 83  46 | 63  48 |
| **Age (years)**  **Median**  **Range** | 55  20-83 | 60  17-88 | 44  20-94 |
| **Cancer Types** |  |  |  |
| Breast | 51 | 8 | - |
| Colorectal | 22 |  | - |
| Lung | 1 | 4 | - |
| Ovary + Peritoneum | 7 | 16 | - |
| Pancreas | 6 | 12 | - |
| Prostate | 9 | 46 | - |
| Head and Neck | 57 | 2 | - |
| Esophagus | 1 |  | - |
| Uterine | 7 |  | - |
| Gastric | 5 |  | - |
| Kidney | 3 | 1 | - |
| Biliary Tract |  | 2 | - |
| Cervix | 4 | 1 | - |
| Thyroid | 5 | 5 | - |
| Liver |  | 1 | - |
| Penis | 3 |  | - |
| Small intestine | 1 |  | - |
| Vulva | 2 |  | - |
| Others | 1 | 31* | - |
| **Total** | **185** | **129** | **111** |

**includes stroke, diabetes, pyrexia, hypertension, infection, or inflammatory conditions.*

**Supplementary Table 3. Demographics of the Low Tumor Burden Study Cohort.**

| **Cohorts** | **Cancer** |
| --- | --- |
| **Gender**  **Male**  **Female** | 27  55 |
| **Age (years)**  **Median**  **Range** | 58  21-82 |
| **Cancer Types** |  |
| Breast | 23 |
| Colorectal | 17 |
| Ovary | 11 |
| Pancreas | 8 |
| Lung | 5 |
| Uterine | 4 |
| Bile Duct | 3 |
| Gastric | 4 |
| Kidney (Renal) | 2 |
| Esophagus | 2 |
| Bladder | 1 |
| Prostate | 1 |
| Thyroid | 1 |
| **Total** | **82** |

**Supplementary Table 4. Demographics of Suspected Pan-Cancer Cases Cohort**

259 individuals suspected of various cancers.

| **Cohorts** | **Initial Suspected** | **Final Diagnosis** | |
| --- | --- | --- | --- |
|  |  | **Cancer** | **Benign** |
| **Gender**  **Male**  **Female** | 92  167 | 80  150 | 11  18 |
| **Age (years)**  **Median**  **Range** | 53  17-92 | 53  19-92 | 50  17-77 |
| **Cancer Types** |  |  |  |
| Breast | 90 | 82 | 8 |
| Colorectal | 12 | 12 | - |
| Lung | 27 | 23 | 4 |
| Ovary + Peritoneum | 30 | 24 | 6 |
| Pancreas | 11 | 11 | - |
| Prostate | 20 | 14 | 6 |
| Head and Neck | 35 | 33 | 2 |
| Esophagus | 1 | 1 | - |
| Uterine | 3 | 3 | - |
| Gastric | 1 | 1 | - |
| Unknown Primary | 3 | 3 | - |
| Kidney | 4 | 4 | - |
| Bladder + Urothelial | 3 | 3 | - |
| Cervix | 11 | 10 | 1 |
| Thyroid | 1 | 1 | - |
| Gallbladder | 4 | 3 | 1 |
| Liver | 2 | 1 | 1 |
| Anal | 1 | 1 | - |
| **Grand Total** | **259** | **230** | **29** |

**Supplementary Table 5. Findings of the Proof of Concept Study Cohort.**

| **Cancer type** | **Positive** |
| --- | --- |
| Breast (n = 685) | 582 (84.96%) |
| Colorectal (n = 357) | 296 (82.91%) |
| Lung (n = 266) | 216 (81.20%) |
| Ovary (n = 171) | 140 (81.87%) |
| Pancreas (n = 159) | 128 (80.50%) |
| Prostate (n = 115) | 97 (84.35%) |
| Head and Neck (n = 109) | 79 (72.48%) |
| Esophagus (n = 76) | 65 (85.53%) |
| Uterine (n = 61) | 49 (80.33%) |
| Gastric (n = 54) | 47 (87.04%) |
| Unknown Primary (n = 42) | 32 (76.19%) |
| Kidney (n = 40) | 36 (90.00%) |
| Biliary Tract (n = 55) | 48 (87.27%) |
| Bladder (n = 35) | 28 (80.00%) |
| Cervix (n = 25) | 23 (92.00%) |
| Thyroid (n = 21) | 20 (95.24%) |
| Other (n = 48) | 39 (81.25%) |
| **Overall (n = 2319)** | **1925 (83.01%)** |

**Supplementary Table 6. Findings of the Case Control Study Cancer Sub-Cohort.** The table presents the CTC detection rate in the 185 cancer cases in the case control study.

| **Cancer Type** | **CTC Positive (n, %)** |
| --- | --- |
| Head and Neck (n = 57) | 48 (84.21%) |
| Breast (n = 51) | 44 (86.27%) |
| Colorectal (n = 22) | 21 (95.45%) |
| Prostate (n = 9) | 9 (100%) |
| Ovary (n = 7) | 6 (85.71%) |
| Uterine (n = 7) | 6 (85.71%) |
| Pancreas (n = 6) | 5 (83.33%) |
| **Others (n = 26)** | **25 (96.15%)** |
| **Overall (n = 185)** | **164 (88.65%)** |

**Supplementary Table 7. Findings of the Cancer cases from the Post-Treatment Low Tumor-Burden Cohort.** The table presents the CTC detection rate in 82 cancer cases with low tumor burden at recurrence.

| **Cancer type** | **CTC Positive (n, %)** |
| --- | --- |
| Breast (n = 23) | 22 (95.65%) |
| Colorectal (n = 17) | 15 (88.24%) |
| Ovary (n = 11) | 9 (81.82%) |
| Pancreas (n = 8) | 8 (100%) |
| Lung (n = 5) | 5 (100%) |
| Uterine (n = 4) | 4 (100%) |
| Bile Duct (n = 3) | 3 (100%) |
| Gastric (n = 4) | 4 (100%) |
| Kidney (n = 2) | 2 (100%) |
| Esophagus (n = 2) | 2 (100%) |
| Bladder (n = 1) | 0 (0%) |
| Prostate (n = 1) | 1 (100%) |
| Thyroid (n = 1) | 1 (100%) |
| **Total (n = 82)** | **76 (92.68%)** |

**Supplementary Table 8. Findings of the Cancer cases from the Prospective Suspected Cases Cohort.** The table presents the CTC detection rate in the 230 cancer cases in the prospective case control study.

| **Cancer type** | **CTC Positive (n, %)** |
| --- | --- |
| Breast (n = 82) | 75 (91.46%) |
| Head and Neck (n = 33) | 21 (63.64%) |
| Ovary (n = 24) | 17 (70.83%) |
| Lung (n = 23) | 17 (73.91%) |
| Prostate (n = 14) | 13 (92.86%) |
| Colorectal (n = 12) | 9 (75.00%) |
| Cervical (n = 10) | 8 (80.00%) |
| Pancreas (n = 11) | 9 (81.82%) |
| Others (n = 21) | 14 (66.67%) |
| **Overall (n = 230)** | **184 (80.00%)** |

**Supplementary Table 9. Reagents and Materials**

| **Item** | **Supplier** |
| --- | --- |
| Recombinant human Anti-CD326 (REA764)-PE-Vio-615 | Miltenyi Biotec |
| Hoechst 33342 (nuclear stain) | Thermo Fisher |
| ProLong Gold Mounting Medium | Invitrogen |
| SKBR3 (Breast Cancer Cell Line) | ATCC |
| MCF7 (Breast Cancer Cell line) | ATCC |
| SW620 (Colon Cancer Cell Line) | ATCC |
| A549 (Lung Cancer Cell Line) | ATCC |
| PL45 (Pancreatic Cancer Cell line) | ATCC |
| OVCAR3 (Ovarian Cancer Cell line) | ATCC |
| PC3 (Prostate Cancer Cell line) | ATCC |
| SJCRH30 (Sarcoma Cell line) | ATCC |
| U87MG (Glioblastoma Cell line) | ATCC |
| A375 (Melanoma Cell line) | ATCC |
